# Preferential expansion upon boosting of cross-reactive “pre-existing” switched memory B cells that recognize the SARS-CoV-2 Omicron variant Spike protein

**DOI:** 10.1101/2021.12.30.21268554

**Authors:** Cory A. Perugino, Hang Liu, Jared Feldman, Blake M. Hauser, Catherine Jacob-Dolan, Anusha Nathan, Zezhou Zhou, Clarety Kaseke, Rhoda Tano-Menka, Matthew A. Getz, Fernando Senjobe, Cristhian Berrios, Onosereme Ofoman, Jacob E. Lemieux, Marcia B. Goldberg, Kerstin Nundel, Ann Moormann, Ann Marshak-Rothstein, John A. Iafrate, Gaurav Gaiha, Richelle Charles, Alejandro B. Balazs, Vivek Naranbhai, Aaron G. Schmidt, Shiv Pillai

## Abstract

In previously unvaccinated and uninfected individuals, non-RBD SARS-CoV-2 spike-specific B cells were prominent in two distinct, durable, resting, cross-reactive, “pre-existing” switched memory B cell compartments. While pre-existing RBD-specific B cells were extremely rare in uninfected and unvaccinated individuals, these two pre-existing switched memory B cell compartments were molded by vaccination and infection to become the primary source of RBD-specific B cells that are triggered by vaccine boosting. The frequency of wild-type RBD-binding memory B cells that cross-react with the Omicron variant RBD did not alter with boosting. In contrast, after a boost, B cells recognizing the full-length Omicron variant spike protein expanded, with pre-existing resting memory B cells differentiating almost quantitatively into effector B cell populations. B cells derived from “ancient” pre-existing memory cells and that recognize the full-length wild-type spike with the highest avidity after boosting are the B cells that also bind the Omicron variant spike protein.

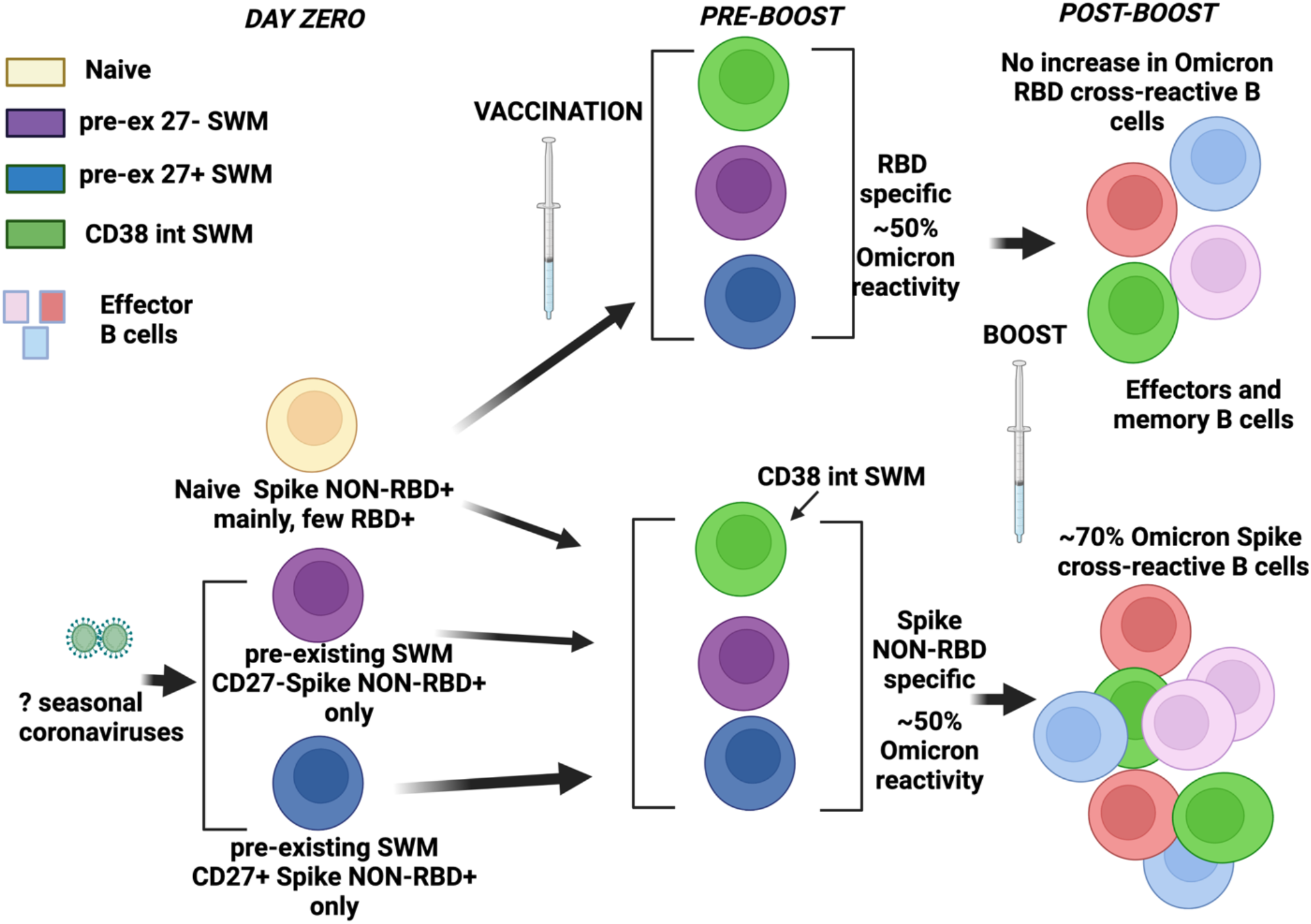

## Introduction

SARS-CoV-2 is a striking example of a novel human pathogen and the recently described SARS CoV-2 Omicron variant (BA.1/B.1.1529) contains novel spike epitopes that have not been encountered before by human B cells. This variant virus can escape neutralization by antibodies in previously vaccinated individuals (Karim and Karim 2021; Cele et al. 2021). The Omicron variant genome contains 59 different mutations including 36 in the gene encoding the Spike protein, of which 15 are in the receptor binding domain (RBD). Booster mRNA vaccination can however induce neutralizing immunity against this variant (Garcia-Beltran et al., 2021).

In humans, switched memory B cells are generally phenotypically defined as IgD-CD27+ B cells and in studies of COVID-19 and mRNA vaccination these cells have often been further categorized based on the expression of IgM, IgG or IgA (Cohen et al.,2021; Kaneko et al., 2020; Rodda et al., 2020; Goel et al, 2021). However, the use of more extensive markers after influenza immunization and the examination of durable antigen-specific B cell populations that are resting or induced has led to the functional description of multiple switched memory B cell subsets including a pre-existing resting switched memory B cell subset (Andrews et al. 2019).

Although there is some sequence homology between the Spike proteins of all seven coronaviruses that infect humans, the RBD region of SARS-CoV-2 has very little similarity to the corresponding domain in other human coronaviruses found in North America (Cueno and Imai, 2021). Consequently, the expectation was that uninfected and unimmunized individuals might harbor pre-existing memory B cells to the SARS-CoV-2 wild type (Wuhan-hu-1) and Omicron variant spike proteins (presumably recognizing partially conserved regions) but likely no pre-existing cross-reactive memory to the wild type or Omicron variant RBDs. We sought to ask how vaccination over an extended period of time and acute and convalescent COVID-19 might differentially influence the evolution and activation of durable pre-existing as well as induced memory B cell populations specific for the wild type and Omicron variant RBD domains.

We show here that resting naive B cells that bind the RBD, including that of wild type SARS-CoV-2, are extremely rare to the point of not being quantifiable, although some RBD binding naïve B cells have been described before using a broader flow cytometric gate (Feldman et al. 2021). Temporal responses to both vaccination and infection revealed three distinguishable antigen-specific durable switched memory B cell compartments; these compartments had expanded and were available for boosting six months after vaccination. These include two pre-existing cross-reactive switched memory B cell compartments. One pre-existing CD27 negative switched memory B cell compartment in unvaccinated and non-infected people contained anti-Spike specific B cells but no RBD binding cells; this compartment as well as a second CD27+ resting pre-existing switched memory B cell compartment, were molded by vaccination and infection to acquire RBD-binding B cells. We show that these pre-existing memory B cell compartments are the main source of cross-reactive Omicron variant RBD-recognizing B cells and that they are activated upon boosting to give rise to induced, activated, durable, switched memory B cell populations as well as to non-durable, transiently activated, antigen-specific B cells. These latter B cells constitute four transiently induced B cell subsets induced by vaccination and/or infection (including two within the canonical IgD^-^CD27^+^ switched “memory” B cell pool) and we categorize these as (non-memory) effector B cell subsets. While only about half of the wild type RBD-binding B cells bound to the Omicron variant RBD, both before and after boosting, boosting preferentially expanded cross-reactive wild type spike binding B cells that also bind the Omicron variant full-length spike protein. The preferential engagement of cross-reactive B cells may represent an evolutionary strategy to deal with variant pathogens such as the Omicron variant of SARS-CoV-2.

## Results

### Heterogeneity of spike-specific B cells following mRNA vaccination

We initially examined fresh blood samples from a small cohort of uninfected and unvaccinated subjects at pre-vaccination (Pre-Vax) and 1-2 weeks post-vaccination following the second dose of an mRNA vaccine (BNT162b2 or mRNA-1273) (Post Vax). We generated two distinct wild type spike-specific tetramer probes, each labeled with a different fluorophore, combined with multi-color flow cytometry to characterize memory and effector B cell subsets (**Figires 1 A, B and C**). **Fig 1 B** illustrates the large number of induced phenotypes that emerge after vaccination. Of the 12 clusters seen using Phenograph, clusters 7 and 9 were further categorized on the basis of being CD27 high or low, yielding a total of 14 clusters **(Figures 1D-E, Table S1)**. Effector B cell subsets and some switched memory B cell subsets could be readily resolved by categorical expression of previously established surface proteins including CD21, CXCR5, CD11c, andCD85j **(Figures 1 C-F)**.

**Figure 1:**
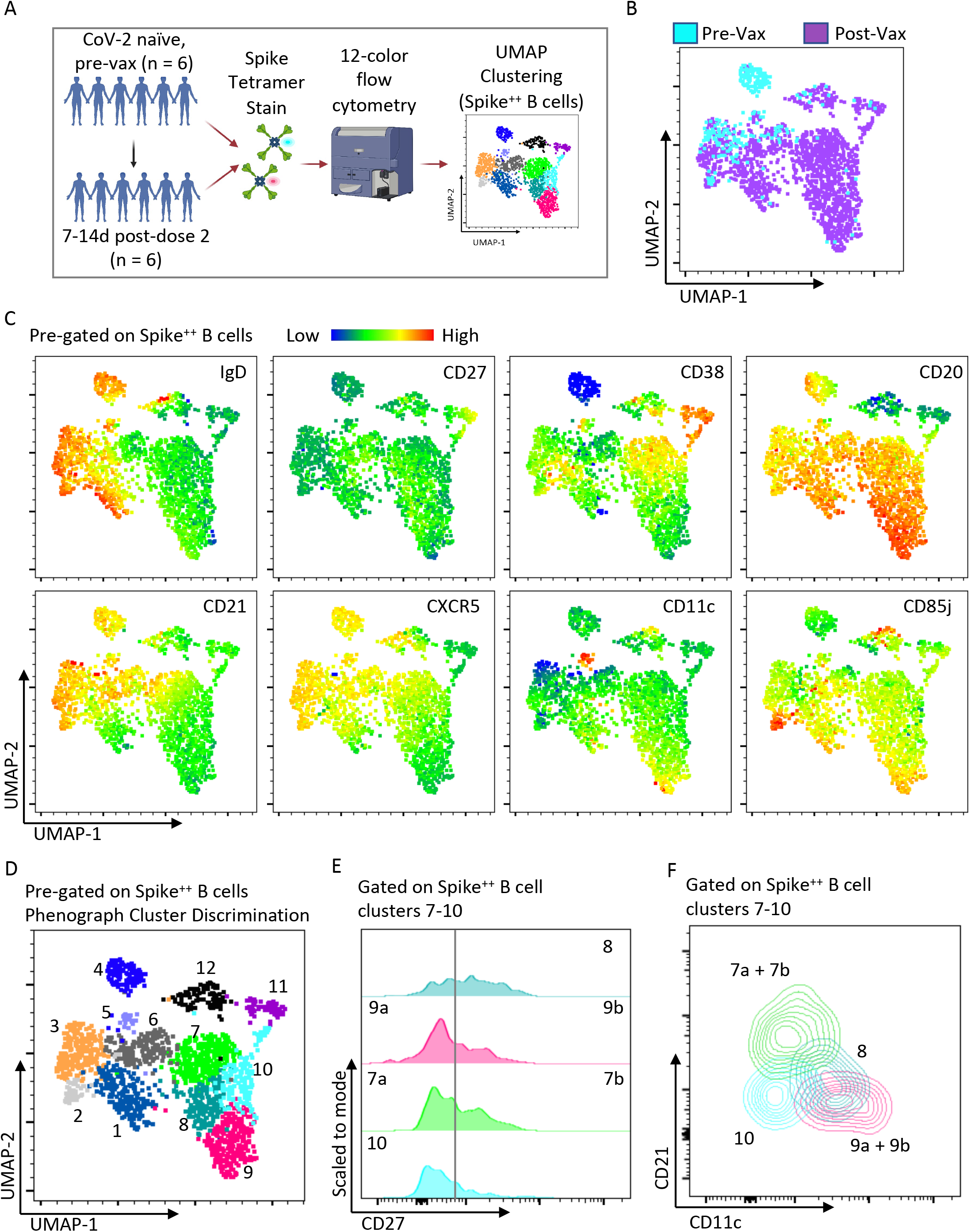
Heterogeneous spike-specific B cells emerge following mRNA vaccination. A) Pictorial overview of initial flow cytometry studies using spike-specific probes and unbiased clustering. B) UMAP dimensionality reduction displaying all Spike-specific B cells from previously unexposed, healthy individuals (n=6) and paired post-vaccination dose #2 (n=6) from the same individuals. Plot is color coordinated based on timepoints of pre-vaccination (Pre-Vax) and post-vaccination (Post-Vax). C) Heatmap overlays of the same UMAP from figure B displaying the relative expression patterns of IgD, CD27, CD38, CD20, CD21, CXCR5, CD11c, and CD85j. D) Overlay of color coordinated clusters among Spike-specific B cells discriminated using Phenograph. E) Histograms displaying relative contributions from double-negative B cells (CD27^Negative^) and switched memory B cells (CD27^+^) among Phenograph clusters 7 through 10. F) Overlaid contour plots of clusters 7 through 10 displaying relative expression of CD21 and CD11c.

### Induced memory and transient effector B cell populations seen after vaccination and infection

Following both mRNA vaccination and natural infection by SARS-CoV-2, we observed the induction of spike-binding circulating B cells that generally persisted proportionally for at least 2-3 months following the completion of the initial vaccination series or into the convalescent phase of COVID-19 **(Figure 2A)**. This corresponded with the induction of a CD27^+^ SWM -type phenotypic change among spike-binding B cells compared to spike-recognizing B cells from pre-vaccination, seronegative donors **(Figure 2B)**. Of the presumed induced switched memory B cell compartments, IgD^-^CD85j^+^CD27^+^ B cells expanded primarily in the context of vaccination and not during acute COVID-19 infection but did persist in the circulation, albeit at a lower frequency in the months following mRNA vaccination **(Figure 2C)**. These cells were also observed among convalescent COVID-19 subjects at a similar frequency to post-vaccination (Vax D36-72) and above the generally absent frequency among pre-vaccinated, seronegative donors **(Figure 2C)**. However as further established below, these cells are not durable, do not persist in the circulation by 6 months following mRNA vaccination, and we have reclassified these as effector, rather than memory, B cells. IgD^-^ CD27^+^ CD85j^-^ CD38^int^ B cells represent the most abundantly expanded, initially induced durable switched memory B cell population both after vaccination and infection **(Figure 2D)**, but two additional more slowly expanding, durable and therefore legitimate, memory B cell compartments are described below in **Figures 3** and **4**.

**Figure 2:**
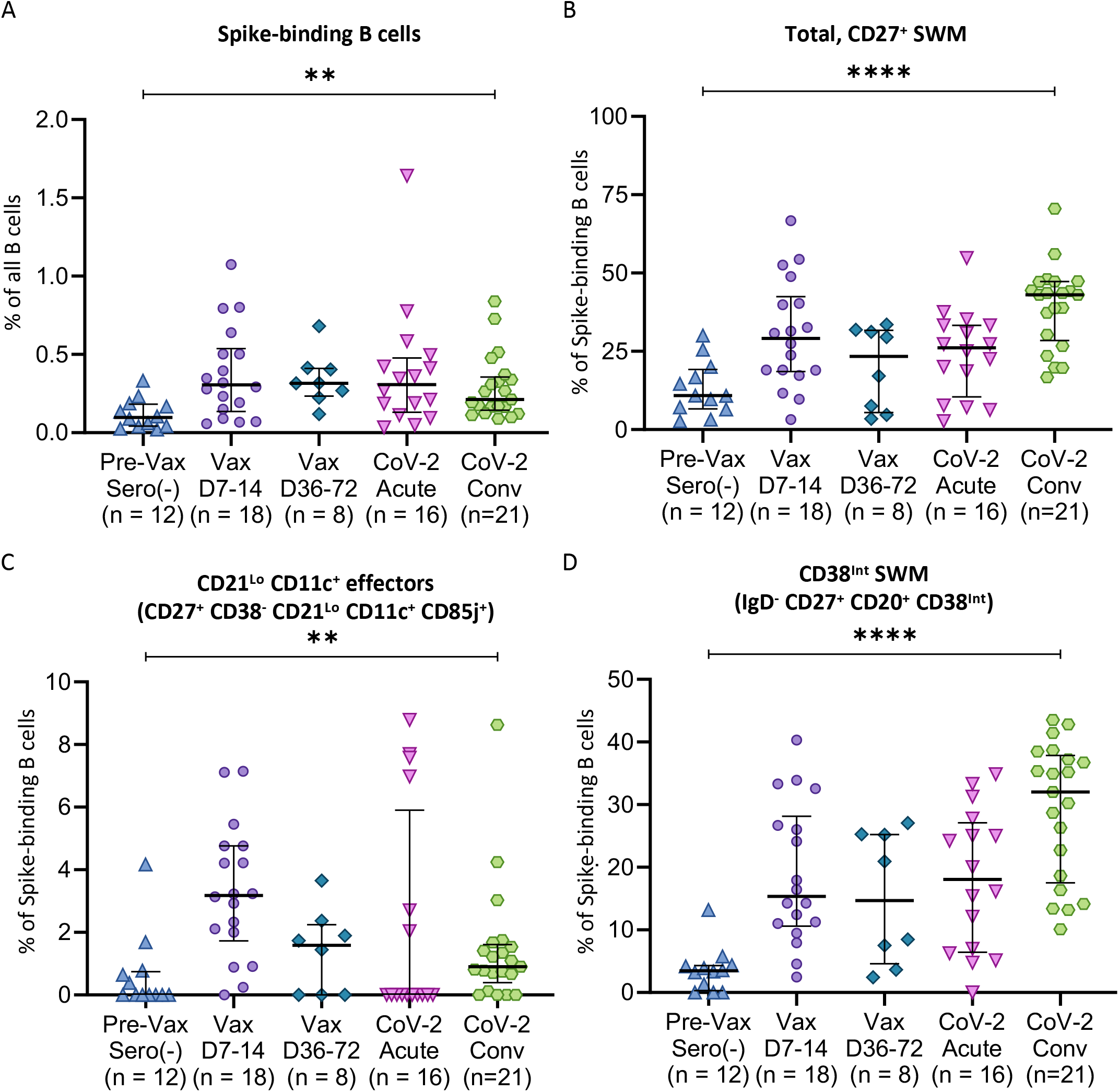
messenger RNA vaccination and acute COVID-19 induce context-dependent, Spike-specific, durable CD27+ SWM B cell responses. Dot plots displaying compartmental effector and memory phenotypic changes of WT Spike-specific B cells among previously unexposed and unvaccinated individuals (n = 12), compared to blood samples from day 7-14 post-vaccination (Vax D7-14, n=18), day 36-72 post-vaccination (Vax D36-72, n=8), acute SARS CoV-2 infected patients (CoV-2 acute, n=16), and convalescent SARS CoV-2 infected patients (CoV-2 Conv, n=21). A) Total Spike-binding B cells as a proportion of total B cells. B) Total CD27+ SWM B cells, C) CD21LoCD11c+ effectors, and D) CD38int memory as proportions of Spike-binding B cells. Statistical differences among cohorts were calculated using the ANOVA test. p-values <0.05 were considered significant. ** indicates p-value 0.001-0.01. **** indicates p-value <0.0001

**Figure 3:**
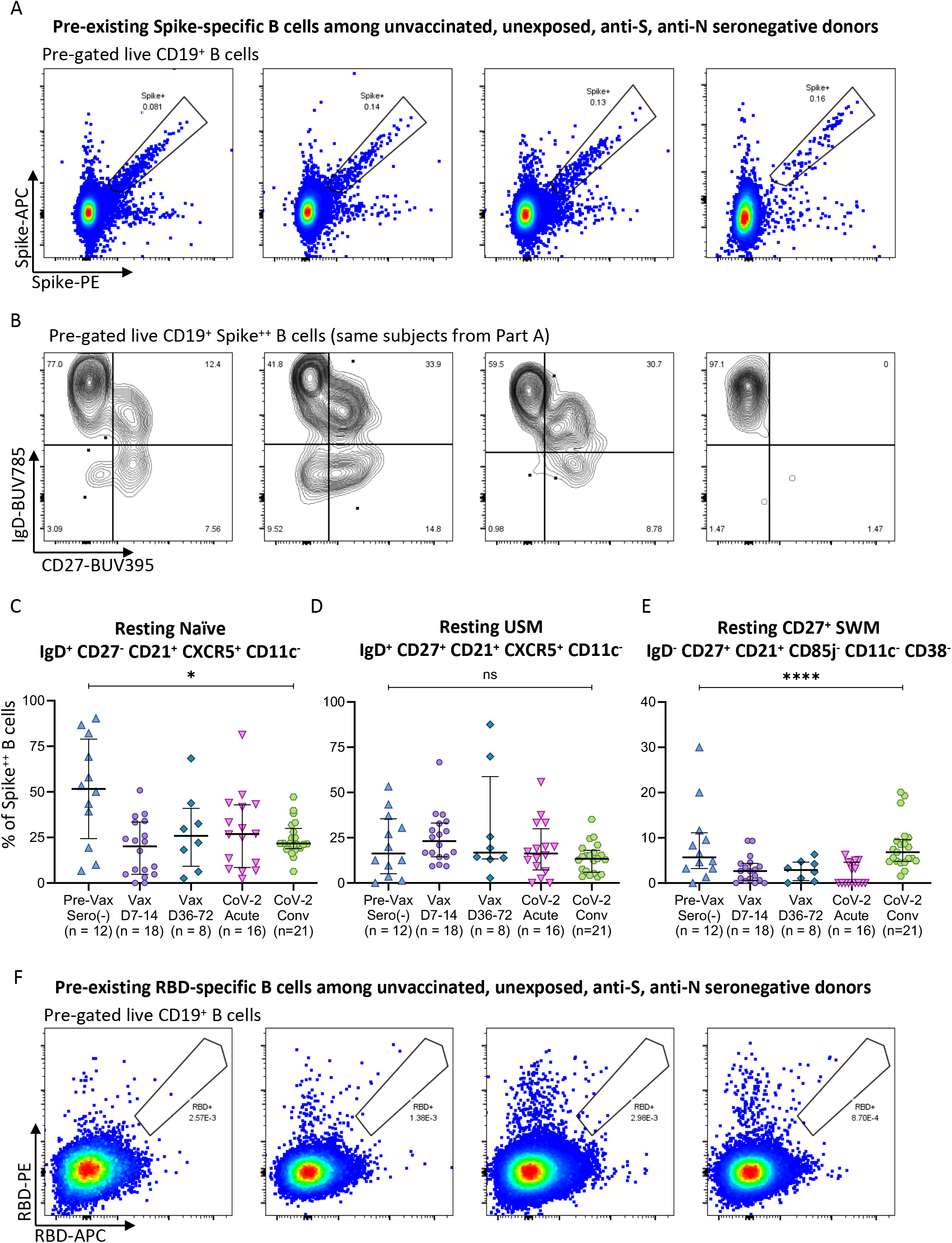
Pre-existing Spike-specific B cell compartments are revealed by profiling unvaccinated, unexposed (SARS-CoV-2 seronegative) donors. A) Flow cytometry pseudo-color plots demonstrating frequent SARS CoV-2 WT Spike-specific B cells among 4 representative previously unexposed, unvaccinated individuals. B) Contour plots displaying standard B cell subsets gating on IgD and CD27 paired with the SARS CoV-2 WT Spike-specific B cells from individuals displayed in part A. C-E) Dot plots displaying composition of pre-existing WT Spike-specific B cells among previously unexposed and unvaccinated individuals (n = 12) compared to blood samples from day 7-14 post-vaccination (Vax D7-14, n=18), day 36-72 post-vaccination (Vax D36-72, n=8), acute COVID-19 patients (CoV-2 acute, n=16), and convalescent COVID-19 patients (CoV-2 Conv, n=21). F) Flow cytometry pseudo-color plots demonstrating extremely rare SARS CoV-2 WT RBD-specific B cells among 4 representative previously unexposed, unvaccinated individuals. Statistical differences among cohorts were calculated using the ANOVA test. p-values <0.05 were considered significant. * indicates p-value 0.01-0.05. **** indicates p-value <0.0001. Key: RBD = receptor binding domain; SWM = switched memory B cells; USM = unswitched memory B cells; ns = not significant.

**Figure 4:**
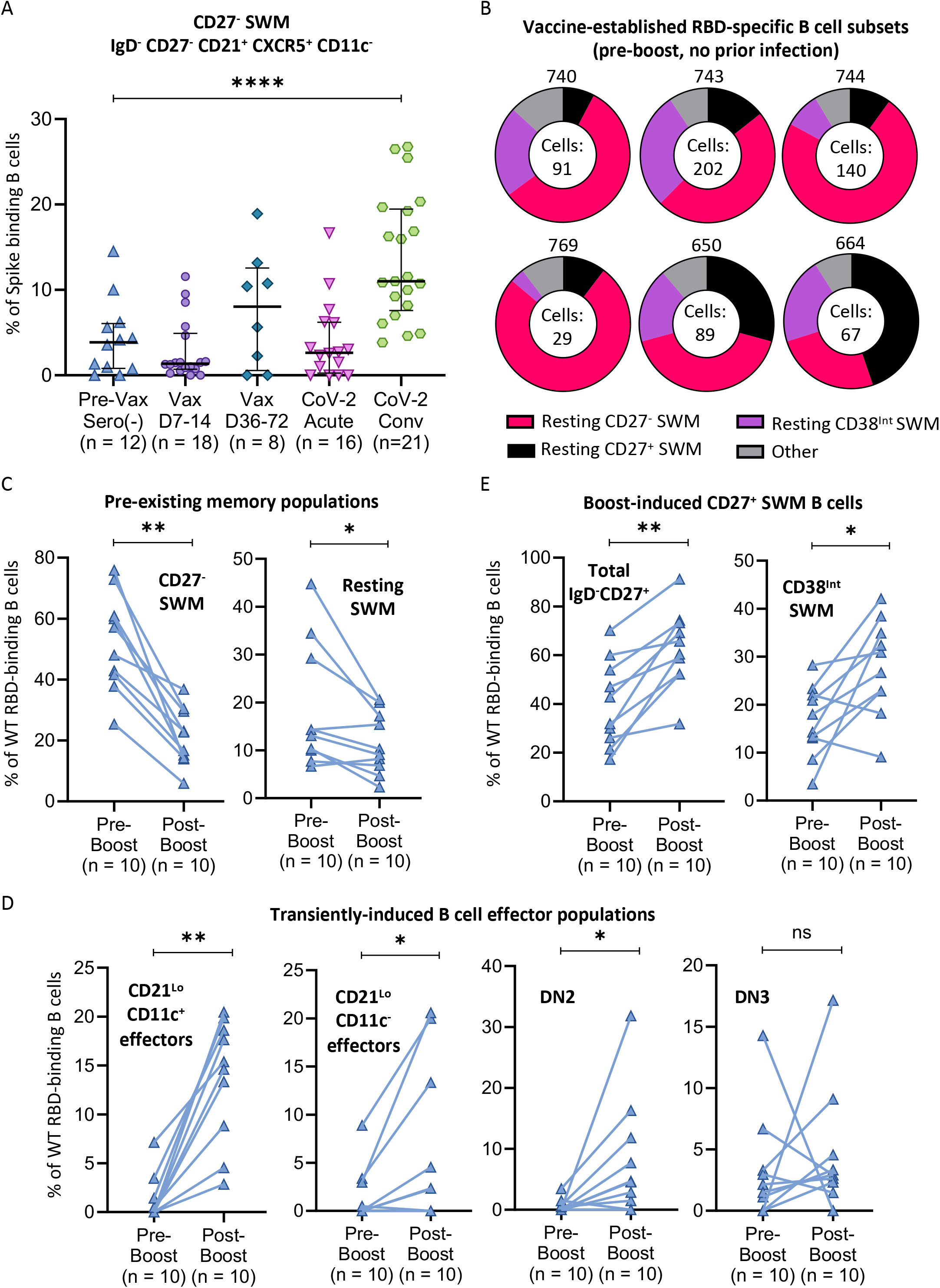
CD27- SWM B cells represent the dominant antigen-specific memory B cell population observed following vaccination and natural infection and give rise to CD27+ memory and transient effector B cell populations upon re-activation. A) Dot plot displaying proportions of WT Spike-specific B cells with a CD27-resting memory phenotype among previously unexposed and unvaccinated individuals (n = 12) compared to blood samples from day 7-14 post-vaccination (Vax D7-14, n=18), day 36-72 post-vaccination (Vax D36-72, n=8), acute COVID-19 patients (CoV-2 acute, n=16), and convalescent COVID-19 patients (CoV-2 Conv, n=21). B) Donut plots representing the cellular distribution of RBD-specific B cell subsets induced by previous vaccination among 6 representative donors pre-boost with no prior COVID-19 infection history. Number of RBD-specific cells analyzed for each subject are included in the donut hole. C-E) Column comparisons of pre-boost (n=10) to paired post-boost (n=10) blood samples comparing C) “pre-existing” memory B cell populations; D) transiently-induced effector B cell populations, and D) vaccine boost-induced CD27+ memory B cell populations. Connecting lines indicate paired samples. Statistical differences among groups were calculated using the ANOVA test or Wilcoxon test, as appropriate. * indicates p-value 0.01-0.05. ** indicates p-value 0.001-0.01. **** indicates p-value <0.0001. Key: RBD = receptor binding domain; SWM = switched memory; ns = not significant

In previous studies, we and others had demonstrated the expansion in the blood of COVID-19 patients of RBD-specific IgD^-^CD27^+^ switched memory B cells, plasmablasts, as well as of IgD^-^CD27^-^CD11c^+^ CXCR5^-^ (DN2, double negative type 2) and IgD^-^CD27^-^CD11c^-^CXCR5^-^ (DN3, double negative type 3) B cells (Kaneko et al., 2020, Woodruff et al., 2020). We asked whether of one or both of the latter populations are induced by COVID-19 vaccination, and potentially represent physiological precursors of plasmablasts or other memory B cells, or if they are disease-specific B cell subsets. As seen in **Fig. S1**, spike-binding DN2 B cells are not seen in unvaccinated and uninfected individuals, transiently expand in most vaccinees soon after vaccination and also in most patients with acute COVID-19 but decline at later time points. These data suggest that DN2 B cells may be induced in the earlier stages of any T-dependent B cell response and though they are prominent in some diseases they are not necessarily disease-specific cells. DN3 B cells on the other hand expand transiently in a small subset of vaccinees but are expanded in most COVID-19 patients and appear to be largely disease-specific B cells. A third population of CD27^+^CD21^lo^CD11c^-^CD38^-^CD85j^-^ B cells, canonically categorized as being within the “switched memory” pool, appears to be a subset of transiently activated switched B cells that are transiently induced after both vaccination and infection do not appear to be true durable memory B cells (**Fig. S1**). The transient nature of these effector “non-memory” B cell populations was more conclusively established at a six-month post vaccination time point when boosting with vaccine re-induced the DN2, CD11c^-^CD85j^-^ and CD11c^+^CD85j^+^ effector B cells that had dwindled with the passage of time. Disease-specific DN3 B cells were not re-induced by boosting (**Figures 4, 5** and **6**).

**Figure 5:**
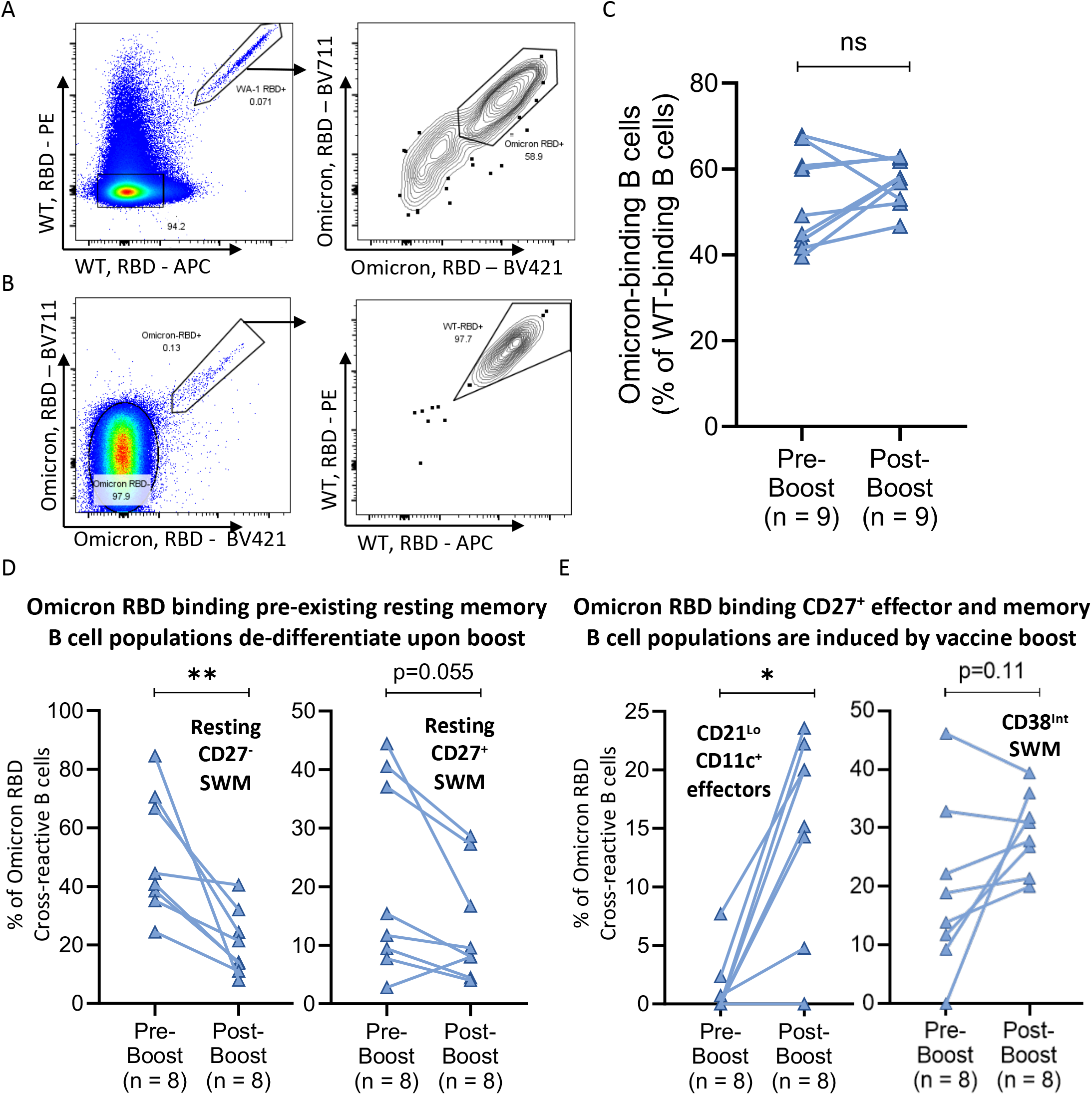
The majority of WT RBD specific B cells can recognize Omicron RBD and mRNA vaccine boost induces resting CD27- and CD27+ memory B cells into CD27+ memory and effector populations. A-B) Flow cytometry pseudo-color and contour plots displaying representative staining of WT-RBD and Omicron-RBD recognizing B cells from a single donor. C-E) Column comparisons of pre-boost (n=10) to paired post-boost (n=10) blood samples comparing C) Omicron-RBD cross-reactivity (as a % of WT-RBD binding B cells); D) Omicron-RBD binding “pre-existing” memory B cell populations; and E) Omicron-RBD binding CD27+ effector and memory B cell populations induced by vaccine boost. Connecting lines indicate paired samples. Statistical differences among groups were calculated using the Wilcoxon test. * indicates p-value 0.01-0.05. ** indicates p-value 0.001-0.01. Key: RBD = receptor binding domain; SWM = switched memory; ns = not significant.

**Figure 6:**
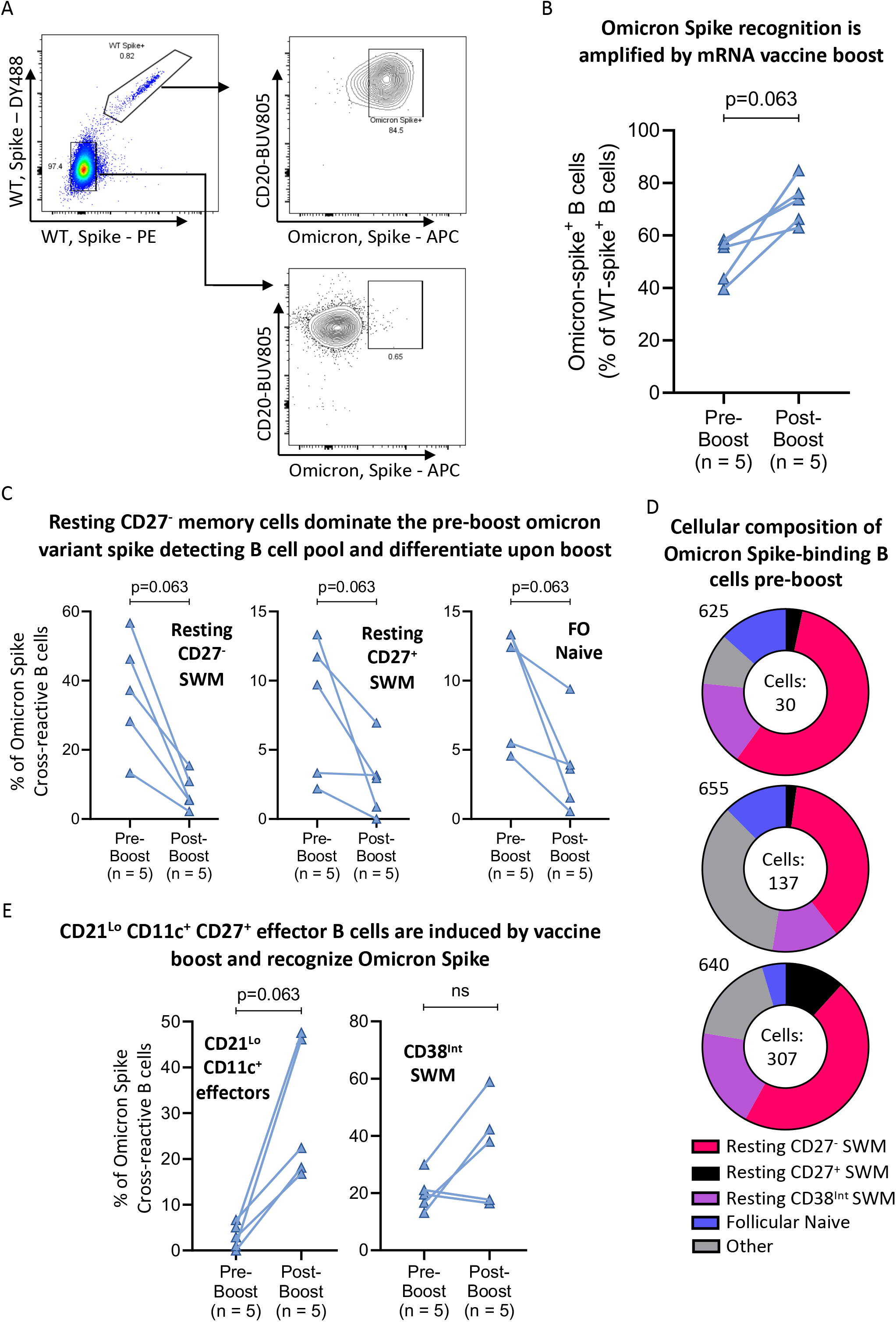
mRNA vaccine boost preferentially expands omicron variant Spike binding B cells and induces resting CD27-memory B cells differentiate into CD21Lo CD11c+ CD27+ effector B cells. A) Flow cytometry pseudo-color and contour plots displaying representative staining of WT-Spike and Omicron-Spike recognizing B cells from a post-boost donor. B, C & E) Column comparisons of pre-boost (n=10) to paired post-boost (n=10) blood samples showing B) the preferential induction of omicron variant spike recognizing B cells by vaccine boost; C) Resting memory and follicular naïve B cells that are induced to differentiate by vaccine boost; and E) Induced CD27+ effector and memory B cell subsets expanded by vaccine boost. Connecting lines indicate paired samples. D) Donut plots representing the cellular distribution of Omicron Spike-binding B cell subsets 6-9 months post-initial vaccination series, pre-boost in 3 representative donors. Number of Omicron Spike-binding cells analyzed for each subject are included in each donut hole. Statistical differences among groups were calculated using the Wilcoxon test. Key: FO = follicular; SWM = switched memory; ns = not significant.

### Pre-existing resting memory B cells that recognize wild type and Omicron variant spike but not wild type (or Omicron variant) RBD

As seen in **Figure 3A**, pre-existing spike binding B cells (for both the wild type and Omicron variant spike protein) were abundant in uninfected, unvaccinated, anti-S negative, anti-N negative subjects. However, wild type (and Omicron variant) RBD-binding B cells are extremely rare in uninfected and unvaccinated subjects (**Figure 3D**). In **Figure 3B**, we show that a large fraction of the spike-binding B cells were found in the relatively broad IgD^+^CD27^-^ “naïve” gate, but spike-binding B cells are also variably present in the unswitched memory population, in the IgD^-^CD27^+^ “switched memory” B cell gate and in the IgD^-^CD27^-^ switched CD27^-^ B cell population **(Figures 3B-E)**. On average a large fraction of wild type and Omicron variant SARS-CoV-2 spike-bonding B cells in uninfected and unvaccinated individuals were naïve follicular B cells **(Figure 3C)**. We included the fourth subject in **Figure 4B** to emphasize that occasional, relatively rare, individuals have no pre-existing cross-reactive memory B cells that recognize the spike protein (suggesting that they were perhaps not adequately exposed to other seasonal coronaviruses).

As seen in **Figure 3C**, after vaccination or infection, the proportion of spike-binding resting naïve follicular B cells (IgD^+^CD27^-^CD21^+^CXCR5^+^CD11c^-^) declined, likely reflecting the expansion of other activated B cells as described below and perhaps some activation of spike-binding naïve follicular B cells in a T-dependent manner. A lower proportion of spike-binding B cells in uninfected and unvaccinated individuals are resting unswitched memory B cells (IgD^+^CD27^+^CD21^+^CXCR5^+^CD11c^-^) which are largely marginal zone B cells - essentially a distinct naïve B cell population - but these cells do not appear to decline with infection and vaccination, suggesting there is very limited or no activation of these spike-binding cells after infection or vaccination **(Figure 3D)**. Some low-level generation of IgM^+^ spike-specific memory B cells after vaccination cannot be ruled out.

A “pre-existing” resting CD27^+^ switched memory B cell compartment (IgD^-^ CD27^+^CD21^+^CD85j^-^ CD11c^-^CD38^-^) also undergoes a relative early decline after acute infection and vaccination and expands again during COVID-19 convalescence **(Figure 3E)**. This population and another resting pre-existing switched memory population described in **Figure 4** assume greater significance when examining the source of Omicron variant and wild type specific RBD and spike responses upon boosting vaccinated subjects, as discussed below.

### CD27-negative pre-existing resting switched memory B cells are the dominant antigen-specific memory B cell population after vaccination or infection

A “pre-existing” resting CD27-negative switched memory B cell compartment (IgD^-^CD27^-^CXCR5^+^CD11c^-^) of spike-binding cells found even in unvaccinated and uninfected seronegative individuals, expands late after vaccination and prominently in COVID-19 patients during convalescence (**Figure 4A**). Among wild type RBD-binding B cells, this existing CD27^-^ resting switched memory B cell compartment (pink in figure) is the dominant, durable antigen-specific memory B cell population in vaccinated individuals six months after vaccination but prior to boosting, with lesser contributions from CD27^+^ resting switched memory B cells (black in figure) and CD38^int^ CD27^+^ memory B cells **(Figure 4B)**. As seen in **Figure 4C**, two of these antigen-specific durable memory B cell compartments decline after boosting with antigen. This observation is much more consistent and striking among CD27^-^ memory B cells. This coincides with an equally striking induction of activated, transiently induced effector B cells including CD21^lo^ CD11c^+^ effectors, CD21^lo^ CD11c^-^ effectors, and DN2 B cells (**Figure 4D**), as well as that of the durable but rapidly expanded CD38^int^ switched memory B cell population (**Figure 4E**). Evidence for the durability of the CD38^int^ population is more apparent when examining B cells that recognize the full-length Omicron variant and wild type spike proteins (**Figure 6**). Six months after vaccination, immunization was seen to have “filled” pre-existing switched memory pools with RBD-binding memory B cells. These cells dominate all RBD-binding B cells in vaccinated individuals **(Figure 4B)**. After boosting, these cells contribute not just to neutralizing antibodies (Garcia-Beltran et al. 2021) that can neutralize the Omicron variant but also to activated effector and rapidly induced switched memory B cell populations.

A similar process occurs for Omicron RBD specific B cells wherein resting pre-existing CD27^+^ and CD27^-^ switched memory B cells dominantly contribute to Omicron RBD binding before the boost and then these very same cells participate dominantly in the expansion of activated omicron specific effector B cells (**Figures 5D-E**). Vaccine boosting appears to effectively induce the differentiation of CD21^lo^CD11c^+^ effector and CD38^int^ memory B cells, seemingly from the resting CD27+ and CD27^-^ switched memory B cell pools. These data establish that pre-existing memory B cells are established by cross-reactivity and represent a key source of humoral immunity to variant pathogen strains.

### Preferential expansion of Omicron variant Spike recognizing B cells after boosting

While about 50% of wild type RBD specific resting memory B cells and virtually no resting naïve follicular B cells recognize the Omicron variant RBD, boosting does not increase the frequency of wild type RBD binding B cells that also recognize the Omicron variant RBD **(Figures 5A-C)**. However, boosting does induce differentiation of pre-existing switched memory B cells into Omicron variant RBD binding effector and memory B cell subsets **(Figure 5E**). In contrast, examination of B cells that bind to full-length wild type and Omicron variant spike proteins reveals that not only do naive B cells also contribute to the initial recognition of Omicron variant full-length spike protein, but vaccine boosting clearly preferentially increases the frequency of wild type spike-binding B cells that also bind the full-length Omicron variant (**Figures 6A-B**), suggesting that the two pre-existing cross-reactive B cell memory populations have evolved to be preferentially engaged by antigens to give rise to effectors **(Figures 6C-E)**. Indeed, while the CD38^int^ memory cell subset expands early and does not appear to differentiate, the two cross-reactive pre-existing memory B cell subsets (resting CD27^+^ and CD27^-^) preferentially differentiate, are phenotypically depleted and are more gradually restored.

Intriguingly, after boosting, a population of wild type spike-binding B cells is easily distinguished as being more avidly decorated by both the labeled wild type spike protein probes (and can therefore be logically inferred to represent B cells with higher affinity B cell receptors for spike). This population is almost entire made up of spike-binding B cells that cross-react with the Omicron variant spike protein (**Fig. S3**).

## Discussion

We describe two pre-existing, cross-reactive, resting switched memory B cell compartments (CD27- and CD27^+^) in unvaccinated and uninfected individuals that already contain memory B cells against the non-RBD portions of the SARS-CoV-2 Wuhan-Hu-1 (wild type) and the Omicron variant spike proteins. In contrast, very few pre-existing memory B cells in uninfected and unvaccinated people recognize the RBD portions of the wild type or Omicron variant spike proteins. It can be surmised that since the spike proteins of seasonal coronaviruses have more homology in the non-RBD portions to SARS-CoV-2 Spike (Cueno and Imai, 2021), pre-pandemic exposure to seasonal coronavirus species may have led to the “filling” of these compartments with anti-spike memory B cells.

With both vaccination and infection, these durable pre-existing resting memory compartments, bereft initially of RBD-specific memory B cells, actively “acquire and hold” wild type and Omicron variant RBD-specific B cells. Strikingly, these compartments account for most RBD binding B cells in vaccinated individuals six months after vaccination. It is these compartments that are targeted by antigen during a boost to rapidly differentiate into effectors. Others have established that boosting also induces neutralizing antibodies (Garcia-Beltran et al., 2021; Doria-Rose et al., 2021) – these are likely generated by preferentially activated cross-reactive B cell clones that recognize conserved epitopes in the SARS CoV-2 Omicron variant Spike protein.

Switched memory phenotype B cells can be germinal center derived, emerging early with little somatic hypermutation perhaps to retain broad specificity to viral variants (Kaji et al. 2012; Viant et al., 2020). However switched memory B cells can also arise in humans in the absence of germinal centers (Kaneko et al. 2020) or in mice in the absence of Bcl-6 (Toyama et al., 2002).

The concept of pre-existing memory has been previously established in influenza immunization-based studies (Andrews et al., 2019). In COVID-19 patients, a single-cell RNA sequence-based study also suggested the existence of some pre-existing memory B cells (Sokal et al., 2021), and indeed there is clearly room for a better understanding of the heterogeneity of functionally relevant and durable memory B cells.

We establish here that SARS CoV-2 infection as well as mRNA vaccination (and vaccine boosting) help distinguish three durable human memory B cell populations that respond to recall antigenic stimuli and bind to wild type and variant antigens with different behaviors and kinetics of expansion. IgD^-^CD27^+^CD38^int^CD11c^-^ switched memory B cells are durable B cells that expand further upon re-exposure/boosting. Resting “pre-existing” CD27+ and CD27-switched memory B cell subsets are activated upon boosting and appear to rapidly differentiate in vivo (with no evidence for initial expansion) in response to antigenic exposure into three distinct transient effector B cell populations, namely IgD^-^CD27^+^CD21^lo^CD11c^+^ switched effector B cells, IgD^-^CD27^+^CD21^lo^CD11c^-^ switched effector B cells and the IgD^-^CD27^-^CXCR5^-^CD11c^+^ DN2 B cell population. A separate transiently generated effector population that does not expand following vaccination or boosting but is seen in acute COVID-19 is the IgD^-^CD27^-^CD11c^-^ CXCR5^-^ DN3 B cell population.

Most interestingly, these studies have provided unexpected information that might yield some potential insights regarding the potential preservation of host immunity against the Omicron variant of SARS-CoV-2 in previously infected or immunized populations. Resting pre-existing memory and naïve follicular B cells that recognize what are likely primarily non-RBD epitopes in the wild type full-length spike protein and that also cross-react with the Omicron variant spike protein preferentially differentiate into effector B cells upon vaccine boosting. This preferential boosting of B cells that recognize cross-reactive B cell epitopes, possibly B cells generated by sequential exposure to seasonal coronaviruses and the wild type SARS-CoV-2 vaccine derived spike protein, might reflect the importance of pre-existing cross-reactive memory in rapid responses to novel pathogens, and may prove key to the possible attenuation of Omicron variant linked COVID-19 disease severity in populations with cross-reactive immunity.

## Data Availability

All data produced in the present study are available upon reasonable request to the corresponding author.

## Acknowledgments

This work was supported by NIH U19 AI110495 to SP and the Massachusetts Consortium on Pathogen Readiness and the UMASS Covid Research Fund. The graphical abstract was prepared using Biorender.

## Author Contributions

Conceptualization (SP, CAP); Methodology (CAP, AGS, ABB); Investigation (CAP, HL); Clinical Resources/patient categorization/sample generation (VN, RC, MBG, JEL, KN, AM-R, AM,GG, JAI, AN, ZZ, CK, RT-M, CB, OO); Reagent generation (AGS, ABB, JF, BMH, CJ-D); Original draft (SP, CAP); Review and editing (SP, CAP, AGS, CJ-D,AM-R, JEL, ABB, MBG, MAG) Supervision (SP).

## Legends to Supplemental Figures

**Figure S1: Effector B cell populations are transiently induced by mRNA vaccination and COVID-19 in a context-dependent manner**. Dot plot displaying proportions of WT Spike-binding effector B cell subsets among previously unexposed and unvaccinated individuals (n = 12) compared to blood samples from day 7-14 post-vaccination (Vax D7-14, n=18), day 36-72 post-vaccination (Vax D36-72, n=8), acute COVID-19 patients (CoV-2 acute, n=16), and convalescent COVID-19 patients (CoV-2 Conv, n=21). Statistical differences among groups were calculated using the ANOVA test. * indicates p-value 0.01-0.05. *** indicates p-value 0.0001-0.001. **** indicates p-value <0.0001. Key: SWM = switched memory; ns = not significant.

**Figure S2: Effector B cell populations are transiently induced by mRNA vaccination and COVID-19 in a context-dependent manner**. Column comparisons of pre-boost (n=10) to paired post-boost (n=10) blood samples analyzing Omicron RBD binding and Omicron-Spike binding CD21Lo CD11c-effector B cells. Connecting lines indicate paired samples. Statistical differences between groups were calculated using the Wilcoxon test. Key: RBD = receptor binding domain; ns = not significant.

**Figure S3: B cells with high affinity for WT Spike are preferentially expanded by mRNA vaccine boost and these cells are more capable of binding viral variants like omicron compared to Spike low affinity B cell receptors**. A-B) Flow cytometry pseudo-color plots from a representative donor displaying the range of mean fluorescent intensity (MFI) for Spike probes observed among Spike-binding B cells before and after mRNA vaccine boost. C) Flow cytometry contour plots displaying differences in Omicron spike binding across WT-Spike negative cells, WT-Spike+ (MFI Low) cells, and WT-Spike+ (MFI High) cells. D) Column comparison of ability to recognize Omicron-Spike across WT-Spike (low MFI) cells and WT-Spike (high MFI) cells. E) Column comparison of pre-boost (n=10) to paired post-boost (n=10) blood samples showing the preferential induction of high affinity WT-Spike binding cells. Connecting lines indicate paired samples. Statistical differences among groups were calculated using the Wilcoxon test. Key: MFI = mean fluorescent intensity. FO = follicular; SWM = switched memory; ns = not significant

## STAR* Methods

### KEY RESOURCES TABLE

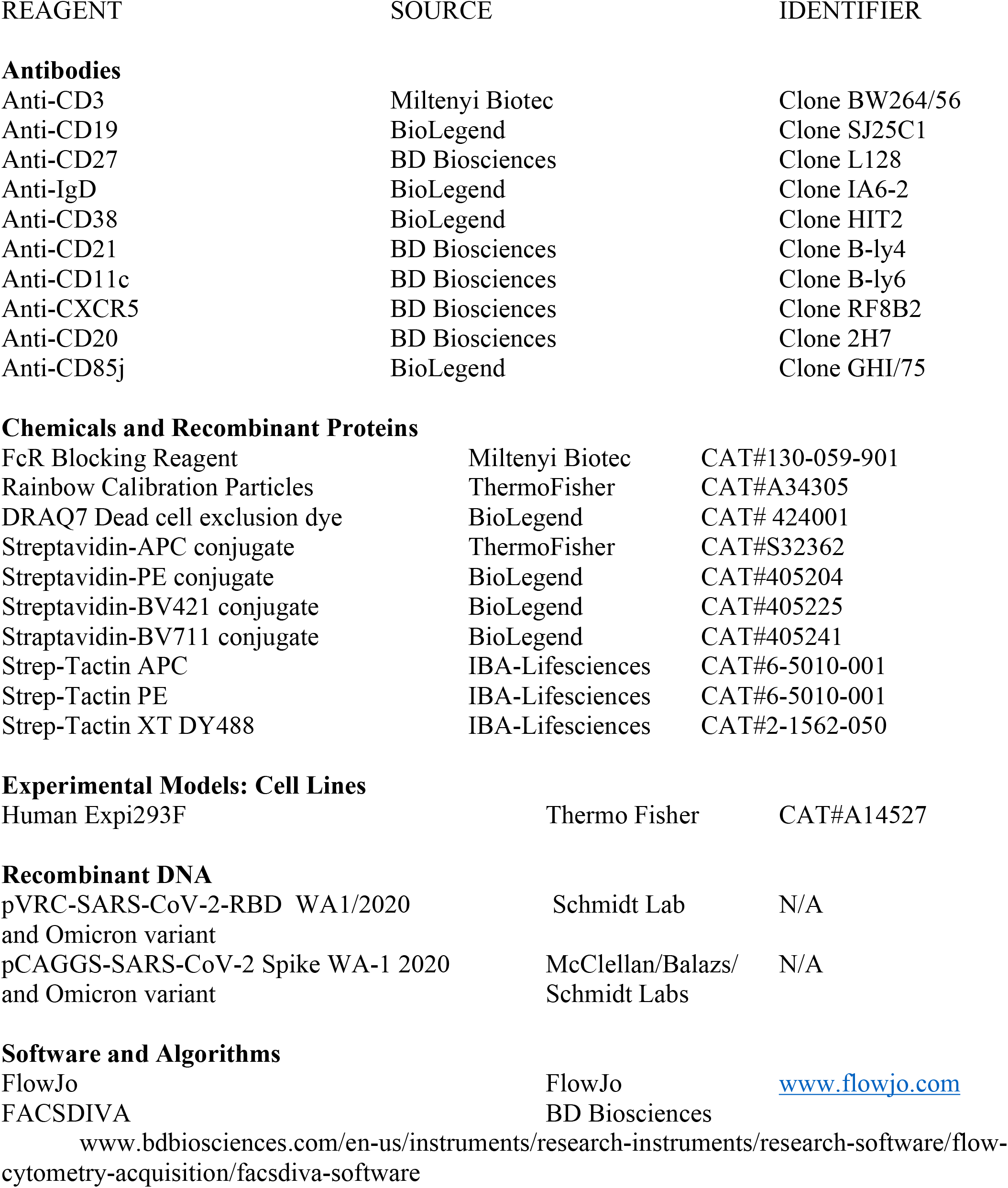
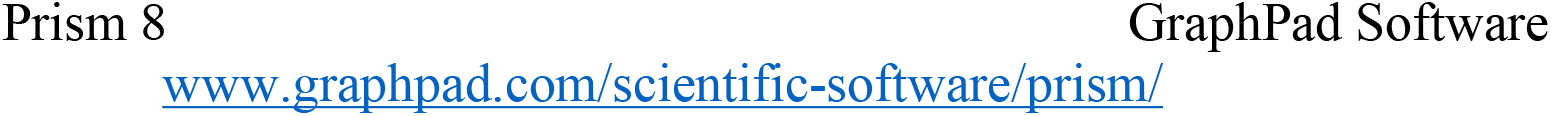

| REAGENT | SOURCE | IDENTIFIER |
| --- | --- | --- |
| **Antibodies** |  |  |
| Anti-CD3 | Miltenyi Biotec | Clone BW264/56 |
| Anti-CD19 | BioLegend | Clone SJ25C1 |
| Anti-CD27 | BD Biosciences | Clone L128 |
| Anti-IgD | BioLegend | Clone IA6-2 |
| Anti-CD38 | BioLegend | Clone HIT2 |
| Anti-CD21 | BD Biosciences | Clone B-ly4 |
| Anti-CD11c | BD Biosciences | Clone B-ly6 |
| Anti-CXCR5 | BD Biosciences | Clone RF8B2 |
| Anti-CD20 | BD Biosciences | Clone 2H7 |
| Anti-CD85j | BioLegend | Clone GHI/75 |
| **Chemicals and Recombinant Proteins** |  |  |
| FcR Blocking Reagent | Miltenyi Biotec | CAT#130-059-901 |
| Rainbow Calibration Particles | ThermoFisher | CAT#A34305 |
| DRAQ7 Dead cell exclusion dye | BioLegend | CAT# 424001 |
| Streptavidin-APC conjugate | ThermoFisher | CAT#S32362 |
| Streptavidin-PE conjugate | BioLegend | CAT#405204 |
| Streptavidin-BV421 conjugate | BioLegend | CAT#405225 |
| Streptavidin-BV711 conjugate | BioLegend | CAT#405241 |
| Strep-Tactin APC | IBA-Lifesciences | CAT#6-5010-001 |
| Strep-Tactin PE | IBA-Lifesciences | CAT#6-5010-001 |
| Strep-Tactin XT DY488 | IBA-Lifesciences | CAT#2-1562-050 |
| **Experimental Models: Cell Lines** |  |  |
| Human Expi293F | Thermo Fisher | CAT#A14527 |
| **Recombinant DNA** |  |  |
| pVRC-SARS-CoV-2-RBD WA1/2020 and Omicron variant | Schmidt Lab | N/A |
| pCAGGS-SARS-CoV-2 Spike WA-1 2020 and Omicron variant | McClellan/Balazs/Schmidt Labs | N/A |
| **Software and Algorithms** |  |  |
| FlowJo | FlowJo | www.flowjo.com |
| FACSDIVA | BD Biosciences |  |
| www.bdbiosciences.com/en-us/instruments/research-instruments/research-software/flow-cytometry-acquisition/facsddiva-software |  |  |

## RESOURCE AVAILABILITY

### Lead Contact

Further information and requests for resources and reagents should be directed to and will be fulfilled by the Lead Contact, Dr. Shiv Pillai

### Materials Availability

The SARS-CoV-2 RBD and Spike proteins are made freely available through Dr. Aaron Schmidt who may be reached through the Lead Contact or directly.

### Data and Code Availability

The published article includes all data generated or analyzed during this study, and summarized in the accompanying tables, figures and Supplemental materials.

## EXPERIMENTAL MODEL AND SUBJECT DETAILS

### Human Subjects

Peripheral blood samples were drawn from patients with COVID-19 at Massachusetts General Hospital and vaccinees at MGH and the University of Massachusetts Medical Center. Freshly isolated PBMCs were used for flow cytometry studies to generate UMAP analyses from Figure 1. Most other PBMCs were stored in liquid nitrogen for subsequent batch experiments.

Sample Size estimation: Power calculations were not undertaken pior to the initiation of these studies. However, the primary end-point was the sum of effector and memory B cells (as a % antigen-specific CD19+ B cells in peripheral blood). Using a two-sided Student’s t-test to compare log values of vaccinated, convalescent and acute COVID-19 patient samples, with groups of 10, we had more than 90% power to detect an effect of size of 1.20 between groups based on simulation studies using 10,000 Monte Carlo samples with a type 1 error rate of 5%.

Allocation to Experimental Groups: Prior exposure to SARS-CoV-2 was determined by measuring serologic responses to SARS-CoV-2 Spike and Nucleocapsid antigens in baseline, pre-vaccination plasma samples. Antibodies to the spike protein receptor binding domain and nucleocapsid were measured by the Roche Elecsys Anti-SARS-CoV-2 S and Anti-N assay (Roche Diagnostics, Indianapolis, IN) per manufacturer’s instructions. Uninfected and unvaccinated subjects were defined by having no detectable anti S and anti-N antibodies.

Convalescence was defined as a clinically asymptomatic state on the date of blood draw, either from a baseline asymptomatic state or recuperated from moderate clinical symptoms of COVID-19. Convalescent COVID-19 patients from a prior asymptomatic state were identified by the presence of Anti-SARS-CoV-2 S or Anti-N responses at the baseline, pre-vaccination time point.

#### Study Approval

This study was performed with the approval of the Institutional Review Boards at the Massachusetts General Hospital and the University of Massachusetts Medical School.

## METHOD DETAILS

### Flow cytometry

Depending on the PBMC yield, a range of 2-10 million fresh PBMCs were stained within 2 hours of isolation for the initial flow cytometry studies. For subsequent batch analyses, cryopreserved PBMCs in aliquots of 5-10 million were thawed quickly at 37°C and rapidly diluted with 1% BSA in PBS. Cells were incubated at 21°C for 15 minutes prior to transfer to FACS tubes for centrifugation. All centrifugation steps were carried out at 1,250 RPM at 4 °C. Prior to antibody staining, Fc receptors were blocked using human FcR blocking reagent (Miltenyi) at a concentration of 1:20 at 4°C for 10 minutes. Strep-tactin or Streptavidin tetramers were generated at the time of each experiment as described below. Cells were surface stained first at 37°C, protected from light, using optimized concentrations of fluorochrome-conjugated primary antibodies for 20 minutes. Cells were then washed, centrifuged, and resuspended in a separate 4°C antibody master mix for an additional 30-minute stain at 4°C. Finally, cells were washed, centrifuged, and re-suspended in DRAQ7 dead cell exclusion dye (BioLegend) at a concentration of 1:1000. Antibodies with manufacturer and clone information is detailed under the STAR Methods section. For the RBD-specific analyses, fluorescently labeled WT-RBD tetramers were generated with Streptavidin-PE (BioLegend) and Streptavidin-APC (ThermoFisher) conjugates while Omicron-RBD tetramers were generated with Streptavidin-BV421 (BioLegend) and Streptavidin-BV711 (BioLegend). For Spike-specific analyses, fluorescently labeled WT-Spike tetramers were generated with Strep-Tactin XT DY488 (IBA) and Strep-Tactin PE (IBA). Omicron-Spike tetramers were generated with Strep-Tactin APC (IBA).

Flow cytometry was performed on a BD Symphony cytometer (BD Biosciences, San Jose, CA) and rainbow tracking beads (8 peaks calibration beads, Fisher) were used to ensure consistent signals between flow cytometry batches.

### RBD and Spike Expression and Purification

SARS-2 RBD (GenBank: MN975262.1), Omicron variant RBD, full-length wild type spike and the Omciron variant RBD were cloned into pVRC vector containing an HRV 3C-cleavable C-terminal SBP-His_8X_ tag and sequence confirmed by Genwiz. The constructs were transiently transfected into mammalian Expi293F suspension cells for recombinant expression. 5 days post-transfection, supernatants were harvested and clarified by low-speed centrifugation. The RBD was purified by immobilized metal affinity chromatography (IMAC) using Cobalt-TALON resin (Takara) followed by size exclusion chromatography on Superdex 200 Increase 10/300 GL (GE Healthcare) in PBS. Purity was assessed by SDS-PAGE analysis. The fluorescent PE-SA (Invitrogen, Cat#12-4317-877) and APC-SA (Invitrogen; Cat#S32362) labels were added to the purified SBP-tagged RBD proteins through iterative complex formation, as previously described (Weaver et al. 2016). The fluorescent SA conjugates were added to SBP-RBD in five increments to sequentially form the complexes. In this case the final molar ratio of probe to streptavidin valency was 1:1 (one SA-fluorophore can bind two SBP tags). After each stepwise addition of the fluorescent label, the mixture was incubated for 20 minutes and set rotating at 4C within an opaque 1.5 mL Eppendorf spin tube. Using this method, we generated fluorescent probes at a final concentration of 0.1ug/ul. An example of SBP-RBD (31,000g/mol) labeled with PE-SA (300,000g/mol) is described for a total 10 assays (0.5 ug labeled protein per assay): First, 5uL of SBP-RBD at 1ug/ul was diluted in 20.8uL PBS. Fluorescent PE-SA at 1ug/ul was then added in five increments, with an incremental volume of 4.8 uL for a final volume of 50uL.

## QUANTIFICATION AND STATISTICAL ANALYSIS

FCS files were analyzed, and B cell subsets were quantified using FlowJo software (version 10). FlowJo plug-ins used for analyses included PhenoGraph and Uniform Manifold Approximation and Projection (UMAP). GraphPad Prism version 8 was used for all statistical analyses and plotting of data points. For comparison of non-parametric and paired samples, the Wilcoxon matched-pairs signed rank test was used. For comparing more than one population, Kruskal-Wallis testing was used with Dunn’s multiple comparison testing.

A p-value of < 0.05 was considered significant.

---

**Table S1:**
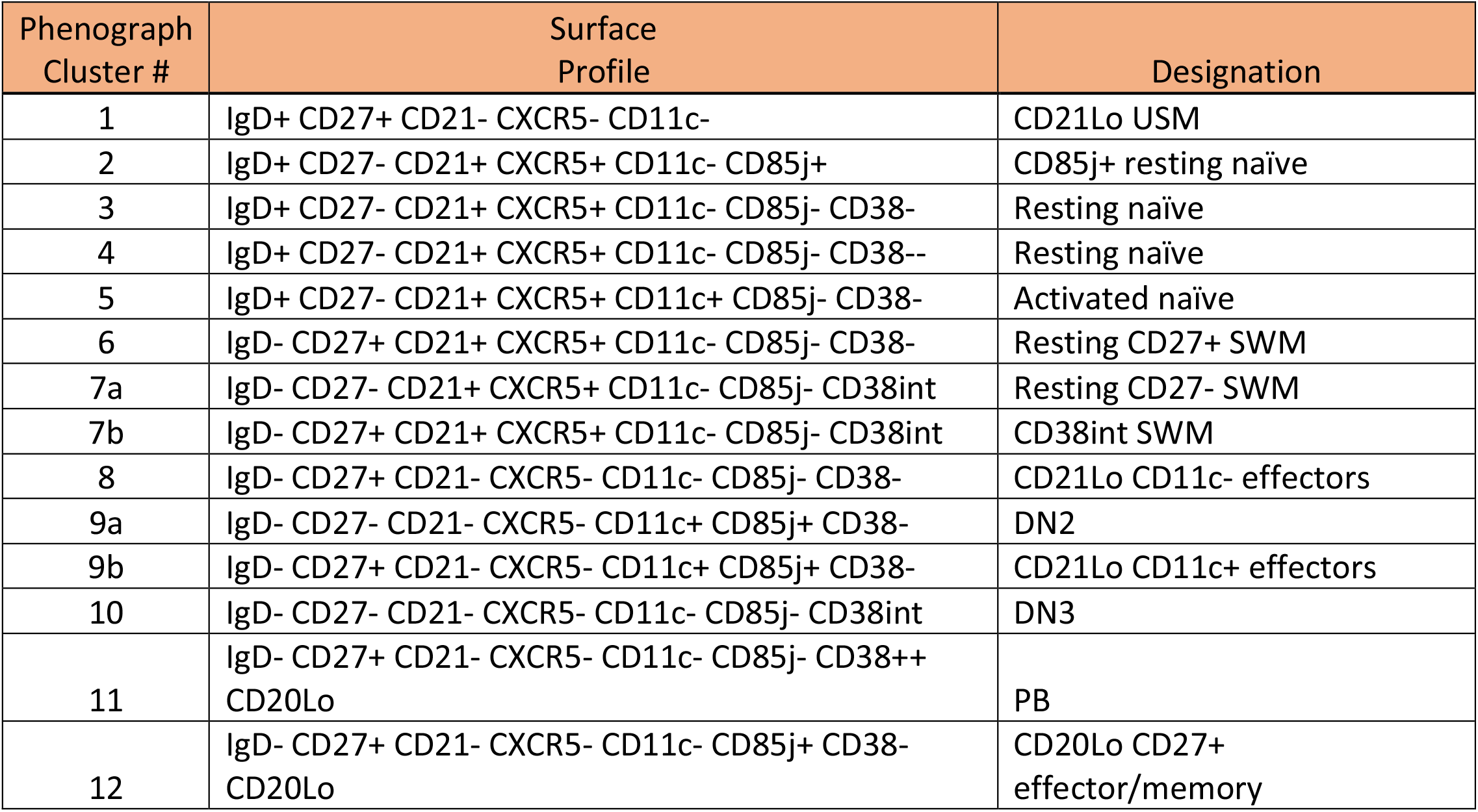
B cell populations based on Phenograph clustering.

