## Supplemental Figures for "Preferential expansion upon boosting of cross-reactive “pre-existing” switched memory B cells that recognize the SARS-CoV-2 Omicron variant Spike protein"

Figure S1

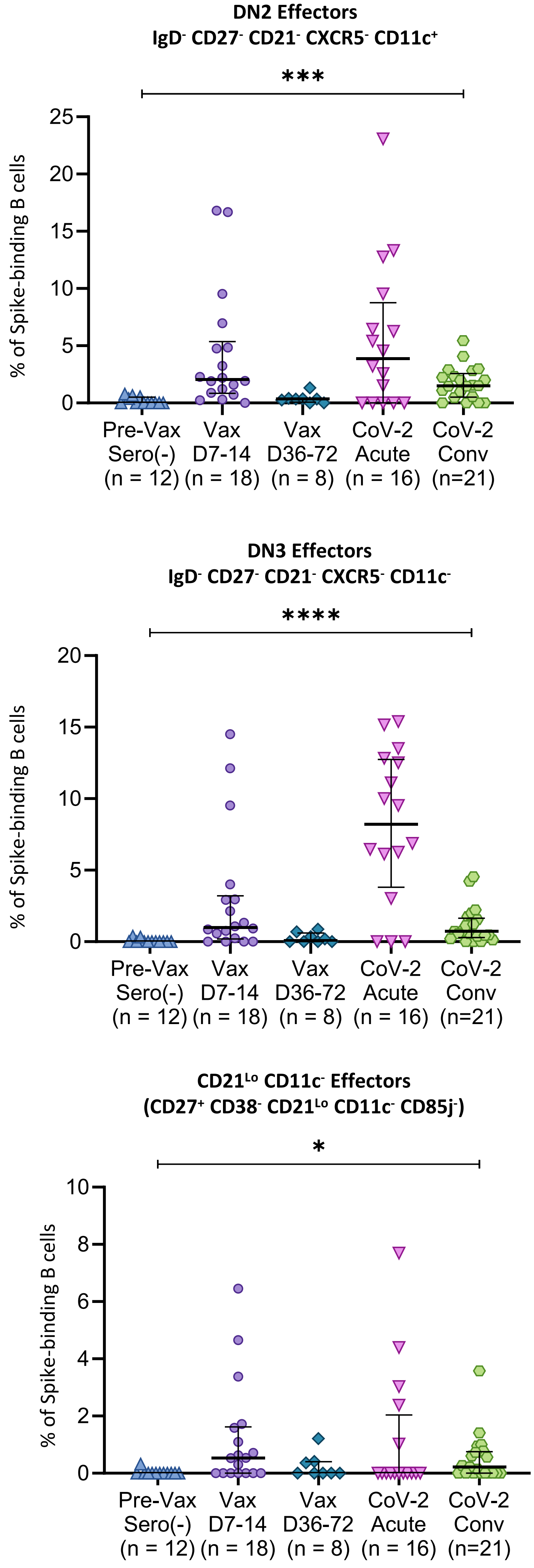


**Figure S1: Effector B cell populations are transiently induced by mRNA vaccination and COVID-19 in a context-dependent manner.** Dot plot displaying proportions of WT Spike-binding effector B cell subsets among previously unexposed and unvaccinated individuals (n = 12) compared to blood samples from day 7-14 post-vaccination (Vax D7-14, n=18), day 36-72 post-vaccination (Vax D36-72, n=8), acute COVID-19 patients (CoV-2 acute, n=16), and convalescent COVID-19 patients (CoV-2 Conv, n=21). Statistical differences among groups were calculated using the ANOVA test. * indicates p-value 0.01-0.05. *** indicates p-value 0.0001-0.001. **** indicates p-value <0.0001. Key: SWM = switched memory; ns = not significant.

### Figure S2

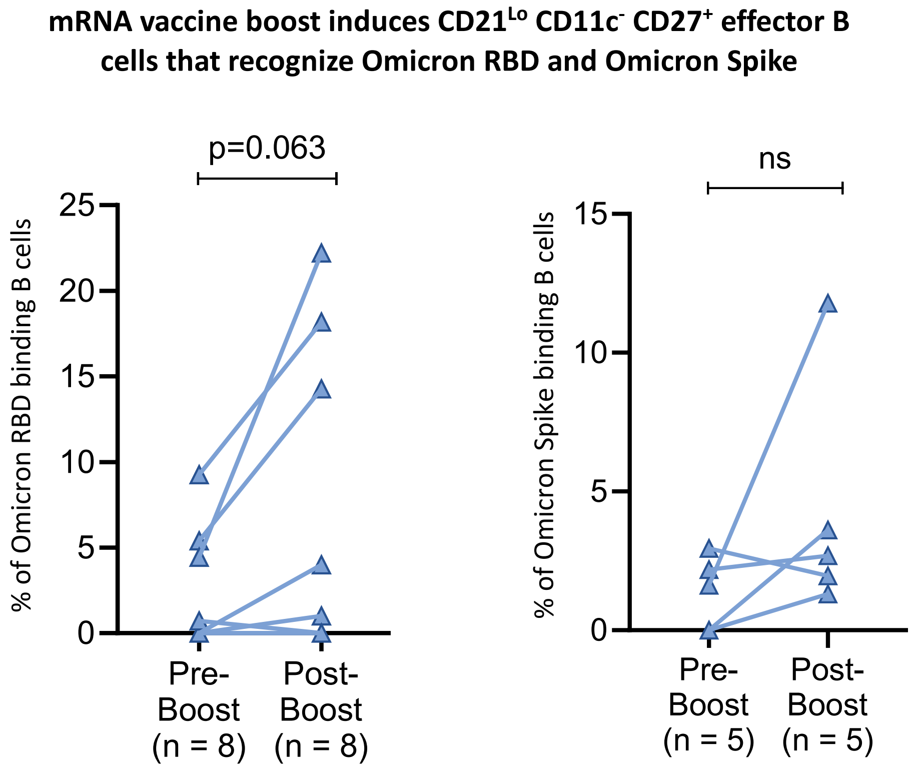


**Figure S2: Effector B cell populations are transiently induced by mRNA vaccination and COVID-19 in a context-dependent manner.** Column comparisons of pre-boost (n=10) to paired post-boost (n=10) blood samples analyzing Omicron RBD binding and Omicron-Spike binding CD21Lo CD11c- effector B cells. Connecting lines indicate paired samples. Statistical differences between groups were calculated using the Wilcoxon test. Key: RBD = receptor binding domain; ns = not significant.

### Figure S3

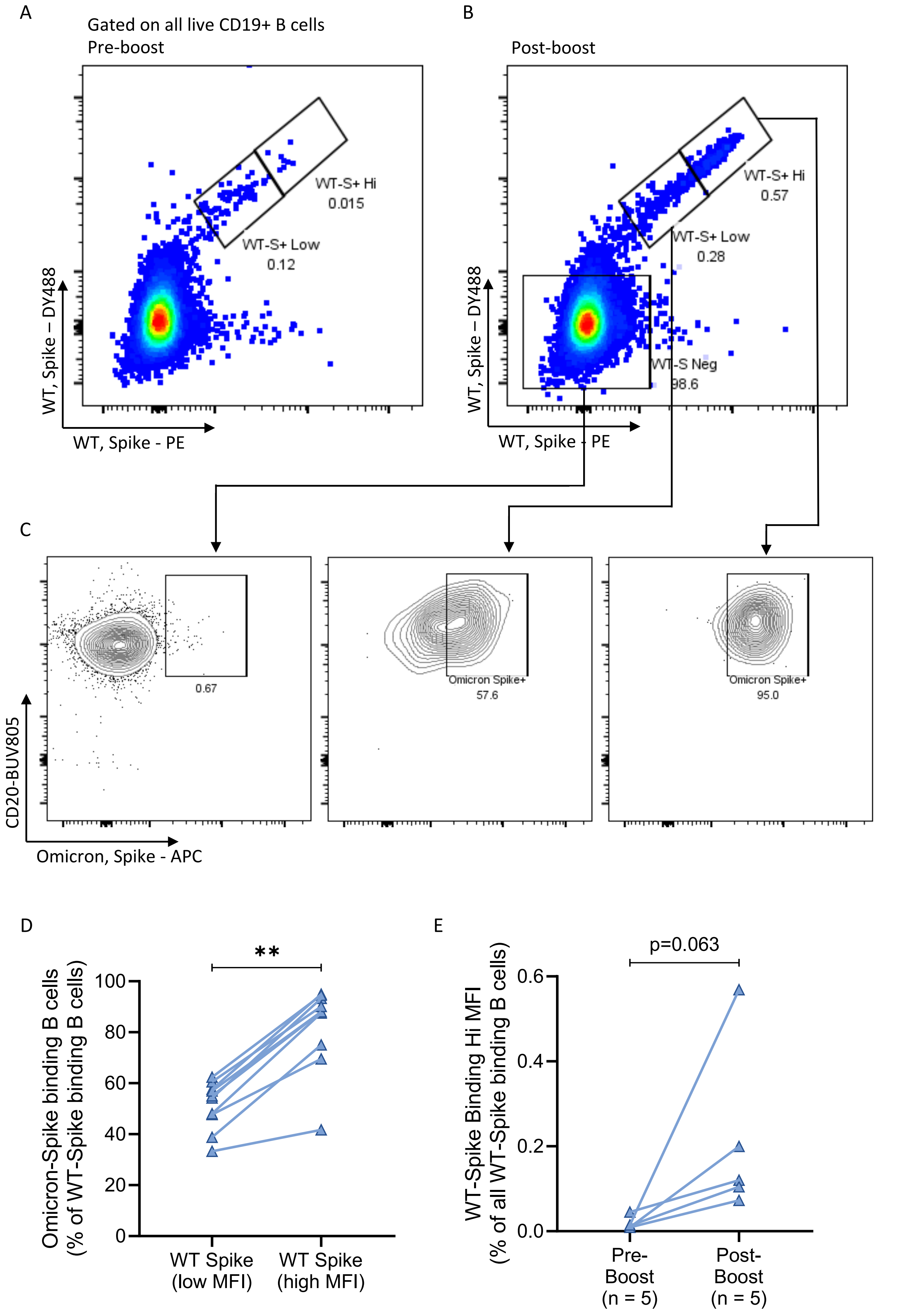


**Figure S3: B cells with high affinity for WT Spike are preferentially expanded by mRNA vaccine boost and these cells are more capable of binding viral variants like omicron compared to Spike low affinity B cell receptors.** A-B) Flow cytometry pseudo-color plots from a representative donor displaying the range of mean fluorescent intensity (MFI) for Spike probes observed among Spike-binding B cells before and after mRNA vaccine boost. C) Flow cytometry contour plots displaying differences in Omicron spike binding across WT-Spike negative cells, WT-Spike+ (MFI Low) cells, and WT-Spike+ (MFI High) cells. D) Column comparison of ability to recognize Omicron-Spike across WT-Spike (low MFI) cells and WT-Spike (high MFI) cells. E) Column comparison of pre-boost (n=10) to paired post-boost (n=10) blood samples showing the preferential induction of high affinity WT-Spike binding cells. Connecting lines indicate paired samples. Statistical differences among groups were calculated using the Wilcoxon test. Key: MFI = mean fluorescent intensity. FO = follicular; SWM = switched memory; ns = not significant
